# Citation reliability of frontier large language models in medical writing and its automated verification

**DOI:** 10.64898/2026.08.31.26361806

**Authors:** Rumi Shin, Jeong-Moo Lee, Jiesuck Park, Ju-Seung Kwun, Hyoung-Won Cho, Si-Hyuck Kang, Ki-Hyun Jeon

**Author notes:** Corresponding author: Ki-Hyun Jeon, MD, Division of Cardiology, Department of Internal Medicine, Seoul National University Bundang Hospital, 82 Gumi-ro 173beon-gil, Bundang-gu, Seongnam 13620, Republic of Korea.

## Abstract

Large language models (LLMs) are increasingly used to draft medical manuscripts, yet their citations are unreliable and clinicians lack a validated way to verify them. We evaluated three frontier LLMs, Claude Opus 4.8, GPT-5.5, and Gemini 3.5 Flash, generating 270 cardiology narrative reviews with web search enabled, and verified all 8,050 references against PubMed. Problematic references accounted for 11.5% of GPT-5.5 output, 29.2% of Claude output, and 29.6% of Gemini output (P < 0.001), with no significant gradient across topics of differing publication volume (P = 0.052). Misattribution, a valid PubMed identifier that resolves to a different article, made up 77% of errors, whereas fabrication was rare (0.6%). Against an expert-adjudicated set of 270 references, an LLM-based Chain-of-Verification (CoVe) detected 60 of 62 problematic references (sensitivity 96.8%, specificity 98.6%), including every misattribution and fabrication. LLM-generated citations require identifier-level verification, and CoVe provides it at expert-level accuracy.

## Introduction

Large language models (LLMs) are increasingly used to draft medical research manuscripts, including the reference lists that accompany them [1,2]. A serious problem is reference hallucination, in which the model produces citations that are fabricated, incomplete, or attributed to the wrong source [3]. Early evaluations of ChatGPT-generated medical content and multidisciplinary literature reviews reported high rates of fabricated or inaccurate references [3,4]. In a systematic-review task, Chelli et al. [5] reported hallucination rates of 39.6% for GPT-3.5, 28.6% for GPT-4, and 91.4% for Bard, then an earlier Google model, indicating that only a small minority of generated references may be fully authentic and accurate in some settings. Similar citation problems have been reported in psychiatry [6], nephrology [7], and internal medicine [8]. A reference hallucination score has since been proposed to quantify the problem across chatbots [9], and further reports cover nursing education [10], stem cell research [11], and trauma education [12]. These findings raise concern about the trustworthiness of scholarship assisted by LLMs.

These estimates, however, predate the current generation of models. Frontier LLMs have advanced rapidly and now operate by default with integrated tooling, most notably web search, so how reliable their citations are in current practice is unclear. Recent work also suggests that citation reliability may vary according to topic familiarity, prompt specificity, and model configuration [13,14]. At the same time, fabricated citations are increasingly recognized as a biomedical publishing integrity problem, with the share of PubMed-indexed papers containing fabricated references rising about 12-fold between 2023 and 2026 [15], and clinicians and editors still lack a validated, practical way to verify the citations that these models produce.

We therefore conducted a study with two aims. The principal aim was to quantify the reference hallucination rate of three current frontier LLMs generating cardiology narrative reviews under their default deployed configuration, and to test whether it depends on the volume of published literature on the topic. The second was to determine how well an LLM-based Chain-of-Verification (CoVe) [16] detects these reference errors, benchmarked against an independent expert gold standard. Because a real citation may still fail to support the sentence attached to it, we also explored whether the same verification approach could be extended from bibliographic accuracy to claim-level support [17,18].

## Methods

### Study design

Each of three frontier models was planned to generate ten independent reviews for each of nine cardiology topics, for 270 narrative reviews in total. Each review was prompted to provide exactly 30 references, giving a planned corpus of 8,100 references. The unit of analysis was the individual reference. The study used no patient data and was not human-subjects research, so institutional review board approval was not required.

### Topics and publication-volume levels

The nine topics were assigned in advance to three publication-volume levels according to the number of PubMed-indexed articles on each topic, used as a measure of how much published literature was available. Counts were measured once on June 18, 2026, using NCBI E-utilities esearch with one fixed query per topic (Supplementary Table 1). The low level comprised transcaval access (N = 191), cardiac contractility modulation (N = 274), and coronary slow flow phenomenon (N = 477). The moderate level comprised subcutaneous implantable cardioverter defibrillator (N = 1,175), bioresorbable vascular scaffold (N = 1,389), and peripartum cardiomyopathy (N = 1,852). The high level comprised sacubitril and valsartan (N = 3,187), left atrial appendage closure (N = 3,780), and cardiac amyloidosis (N = 4,849). Topics were deliberately diverse across procedures, devices, diseases, and drugs so that publication volume would not be confounded with a single topic type.

### Models and generation

Three frontier models were evaluated via their application programming interfaces (APIs) with each provider’s native web-search or grounding tool enabled. These were Claude Opus 4.8 (Anthropic), GPT-5.5 (OpenAI), and Gemini 3.5 Flash (Google). No PubMed-specific retrieval tool was provided during generation. Each review was generated in an independent session with an identical prompt (Supplementary Table 2) that requested a narrative review of 600 to 900 words with exactly 30 Vancouver-style citations, each reference reported as structured JSON with author, title, journal, year, volume, issue, pages, PMID, and DOI. Sampling parameters such as temperature were left at provider defaults and recorded. The exact model string returned by each API response was logged.

### Automated PubMed-based field verification

For the primary model comparison, each stated reference was checked automatically against the bibliographic record in PubMed. The check was a fixed rule-based program and did not use an LLM. It therefore applied the same criteria to all model outputs.

For each reference, the check used three steps. First, it resolved the stated PMID, or mapped the stated DOI to a PMID, and retrieved the PubMed record. Second, it searched PubMed using the stated title and first-author surname, which tested whether the stated identifier actually belonged to the article named by the model. Third, it compared the stated bibliographic fields with the retrieved record under a prespecified rubric that tolerated benign discrepancies, such as a one-year difference between electronic and print publication dates (Supplementary Table 3). Crossref was used only to avoid labeling a real article outside PubMed as fabricated.

Each reference was assigned to one of seven categories, verified, verified benign, real but not in PubMed, field error (correct article with at least one non-benign field wrong), misattribution (the stated identifier pointed to a different article, or the named article carried an incorrect identifier), fabrication (no such article found), and omission (no identifier stated), with full definitions in Supplementary Table 3. The primary outcome was the proportion of problematic references, defined as field error, misattribution, or fabrication, among references with a stated identifier. Benign discrepancies were excluded from the numerator and reported separately.

### Expert validation set

To validate automated verification independently, 270 references were sampled by stratifying the 270 reviews on model crossed with hallucination-rate tercile, giving nine cells with one review per cell. A single expert cardiologist adjudicated each reference, blinded to model identity, as real, field error, misattribution, or fabricated, using all available sources (PubMed, Crossref, Google Scholar). This adjudicated set served as the gold standard. The same set was used to estimate the accuracy of the automated field check and, by Rogan-Gladen correction, to test whether plausible misclassification by that check changed the model ranking (Supplementary Tables 6 and 7).

### Chain-of-Verification implementation

The primary question in this arm was how well an LLM-based Chain-of-Verification (CoVe) detects problematic references. CoVe was introduced by Dhuliawala et al. [16] as a general method for reducing hallucination in LLM output. The original method has four steps. The model first drafts a baseline response. It then plans a set of verification questions about the factual claims in that draft. Each question is answered separately, and in the factored variant the model answers each question without seeing the baseline draft, so an error in the draft cannot propagate into its own verification. The answers are then cross-checked against the draft to produce a final verified response. In the original work the factored variant outperformed joint verification, and open questions outperformed yes-or-no confirmation questions.

We adapted this structure to reference verification with four working principles. First, factored execution. Each verification question was answered in an independent tool call, and the stated reference text served only as the query target, never as evidence. Second, tool grounding. The answer to every question was the value returned by a PubMed tool call, not the model’s own knowledge, because the ground truth for a citation lives in the bibliographic database rather than in the model. Third, open questions. The procedure asked what the record contains rather than whether a stated value is correct. Fourth, atomic decomposition. A reference is a linked set of small claims, so it was verified as five separate questions rather than as one judgment. The five questions covered the resolution of the stated identifier, a reverse search of the stated title and first author, and the authors, journal and year, and volume and pages of the confirmed record (Supplementary Table 8).

The first two questions together establish the identity of the cited article, and the remaining three test its bibliographic fields. When the stated identifier resolved to the article named by the model, the reference was confirmed and the field questions determined whether it was verified or carried a field error. When the stated identifier resolved to a different real article and the reverse search recovered the named article, the reference was classified as a misattribution. When neither step could locate the named article in PubMed or Crossref, the reference was classified as a fabrication (Supplementary Table 9). The same benign tolerances applied as in the automated field check. CoVe was run once on the 270-reference validation set, processed review by review in batches of 30 references, blinded to the expert gold standard, and without tuning to the validation set. Against the expert gold standard we report CoVe sensitivity and specificity overall and its detection rate within each error type.

### Claim-level verification demonstration

As an exploratory extension, we demonstrated claim-level verification, in which the abstract of the cited article is checked against the statement it supports in the review body. From each of the nine validation reviews we selected one reference that both verification methods had classified as verified and that was cited with a substantive factual or quantitative statement. The in-text sentence carrying the citation was extracted as the claim, the cited abstract was retrieved from PubMed, and a single reviewer judged, using the abstract as the only source of truth, whether it supported the claim. Selection was fixed in advance and blind to whether the claim would hold (Supplementary Note 1, Supplementary Table 10).

### Statistics and reproducibility

The primary outcome was modeled with mixed-effects logistic regression on 8,036 references with a stated identifier. The model included LLM, publication-volume level, and their interaction as fixed effects, with random intercepts for topic (n = 9) and review (n = 270) to account for the nesting of references within reviews and topics. Overall effects of LLM, publication-volume level, and their interaction were tested with likelihood-ratio tests comparing nested models, reported as χ² statistics with their degrees of freedom. Pairwise comparisons between models were z tests on the log-odds scale with Bonferroni correction for three comparisons. A prespecified ordered linear contrast tested whether problematic-reference rates decreased from low to moderate to high publication-volume levels. All tests were two-sided with α = 0.05, and exact P values are reported. Adjusted probabilities are reported with 95% confidence intervals. CoVe performance was summarized as sensitivity and specificity against the expert gold standard with Wilson 95% confidence intervals. A supplementary sensitivity analysis applied the Rogan-Gladen correction with review-clustered bootstrap confidence intervals (3,000 resamples) to assess whether possible misclassification by the automated check changed the model ranking. Mixed-effects models and adjusted estimates were computed in R 4.4.0 using lme4 (version 2.0.1) and emmeans (version 2.0.2), and reference verification and supplementary analyses were performed in Python 3.12.2 with NumPy 2.4.1.

## Results

### Reference hallucination according to model

The study design specified 270 narrative reviews with 30 requested references each across nine cardiology topics (Table 1). After parsing model outputs, 8,050 references were available for verification. Fourteen references lacked a stated PubMed identifier (PMID) or digital object identifier (DOI) and were classified as omissions, leaving 8,036 references with a stated identifier for the primary analysis.

**Table 1.** Review topics and generated references.

| **Publication-**<br>**volume level** | **Topic** | **Topic type** | **PubMed-**<br>**indexed articles** | **Reviews**<br>**generated** | **References**<br>**available for**<br>**verification** | **References**<br>**with stated**<br>**identifier** |
| --- | --- | --- | --- | --- | --- | --- |
| Low | Transcaval access | Procedure | 191 | 30 | 900 | 891 |
| Low | Cardiac contractility modulation | Device therapy | 274 | 30 | 900 | 897 |
| Low | Coronary slow flow phenomenon | Disease or<br>phenomenon | 477 | 30 | 902 | 902 |
| Moderate | Subcutaneous implantable<br>cardioverter defibrillator | Device | 1,175 | 30 | 900 | 900 |
| Moderate | Bioresorbable vascular scaffold | Device or stent<br>technology | 1,389 | 30 | 900 | 900 |
| Moderate | Peripartum cardiomyopathy | Disease | 1,852 | 30 | 900 | 899 |
| High | Sacubitril and valsartan | Drug therapy | 3,187 | 30 | 890 | 889 |
| High | Left atrial appendage closure | Procedure | 3,780 | 30 | 868 | 868 |
| High | Cardiac amyloidosis | Disease | 4,849 | 30 | 890 | 890 |

Across these 8,036 references, the crude proportion of problematic references was 11.5% for GPT-5.5, 29.2% for Claude Opus 4.8, and 29.6% for Gemini 3.5 Flash. The proportion of problematic references differed across the three models (likelihood-ratio χ^2^ [df = 2] = 109.5, P < 0.001). GPT-5.5 had about one third the odds of a problematic reference compared with Claude and Gemini (odds ratio [OR] 0.295 vs Claude and OR 0.297 vs Gemini, both Bonferroni-adjusted P < 0.001), whereas Claude and Gemini did not differ (OR 1.01, P = 1.000). The distribution of per-review problematic reference rates by model and publication-volume level is shown in Fig. 1.

**Figure 1.**
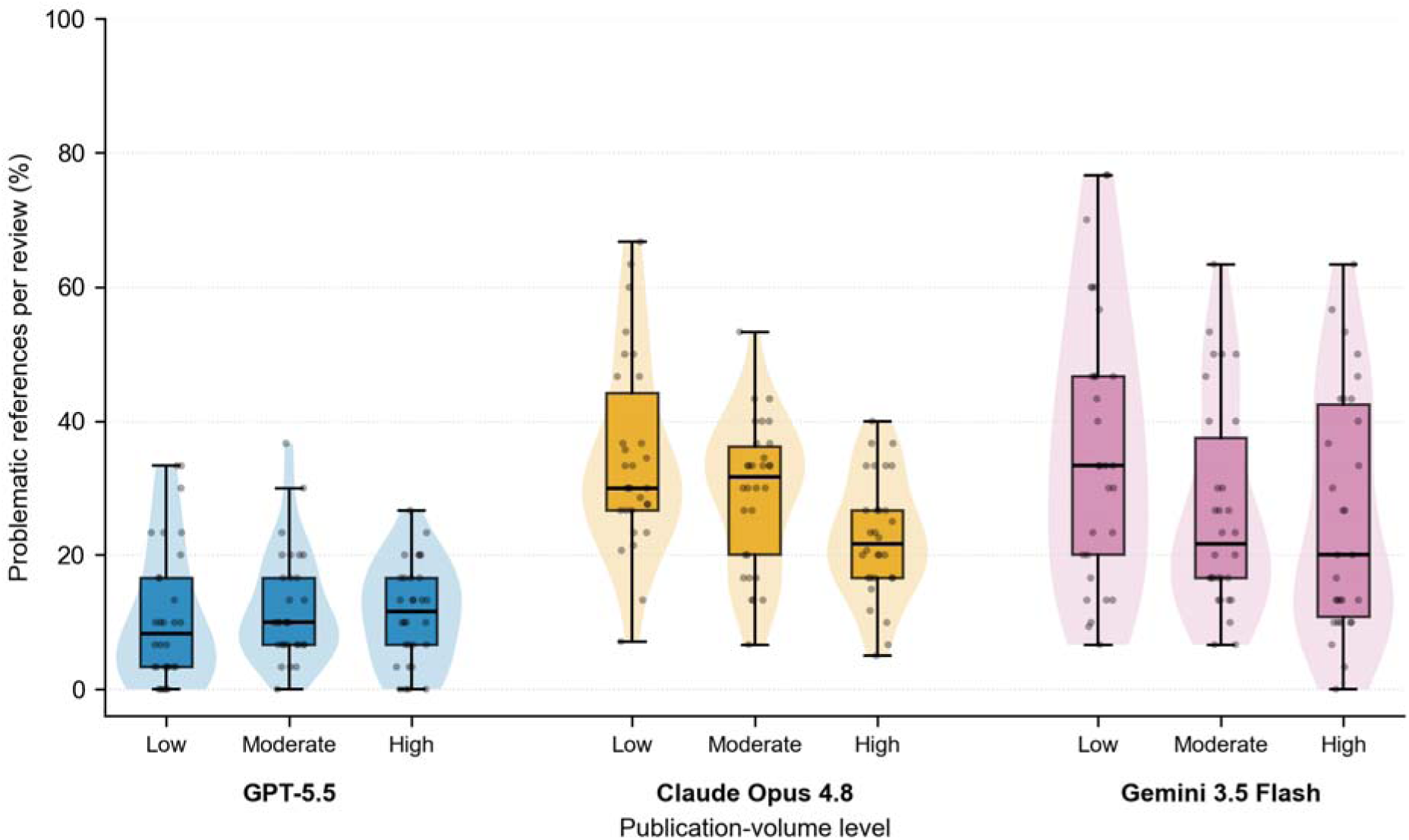
Per-review problematic reference rate by model and publication-volume level. Each point is one generated review (n = 30 reviews per model and level, 270 in total). The rate is the proportion of references with a stated identifier that were classified as problematic (field error, misattribution, or fabrication) by the automated PubMed-based check.

### Publication volume and hallucination

Problematic reference rates were numerically lower in topics with higher publication volume, but the overall publication-volume effect was not statistically significant (likelihood-ratio χ^2^ [df = 2] = 3.77, P = 0.152). The prespecified ordered contrast showed a decreasing pattern from low to moderate to high publication-volume levels, although it was not statistically significant (estimate −0.361 on the log-odds scale, P = 0.052). The interaction between model and publication-volume level was also not statistically significant (χ^2^ [df = 2] = 8.35, P = 0.080).

### Reference error types by model

Misattribution was the dominant error type across models, accounting for 76.8% of all problematic references (Fig. 2, Table 2). Field errors accounted for 22.6% of problematic references and included incorrect bibliographic details such as DOI, author, journal, year, volume, issue, or pages. Fabrication, in which no corresponding article was found, was uncommon and accounted for 0.6% of problematic references. Most erroneous citations were therefore superficially credible. The identifier often existed and resolved, but to the wrong article, or the article was correct but one or more bibliographic fields were wrong.

**Figure 2.**
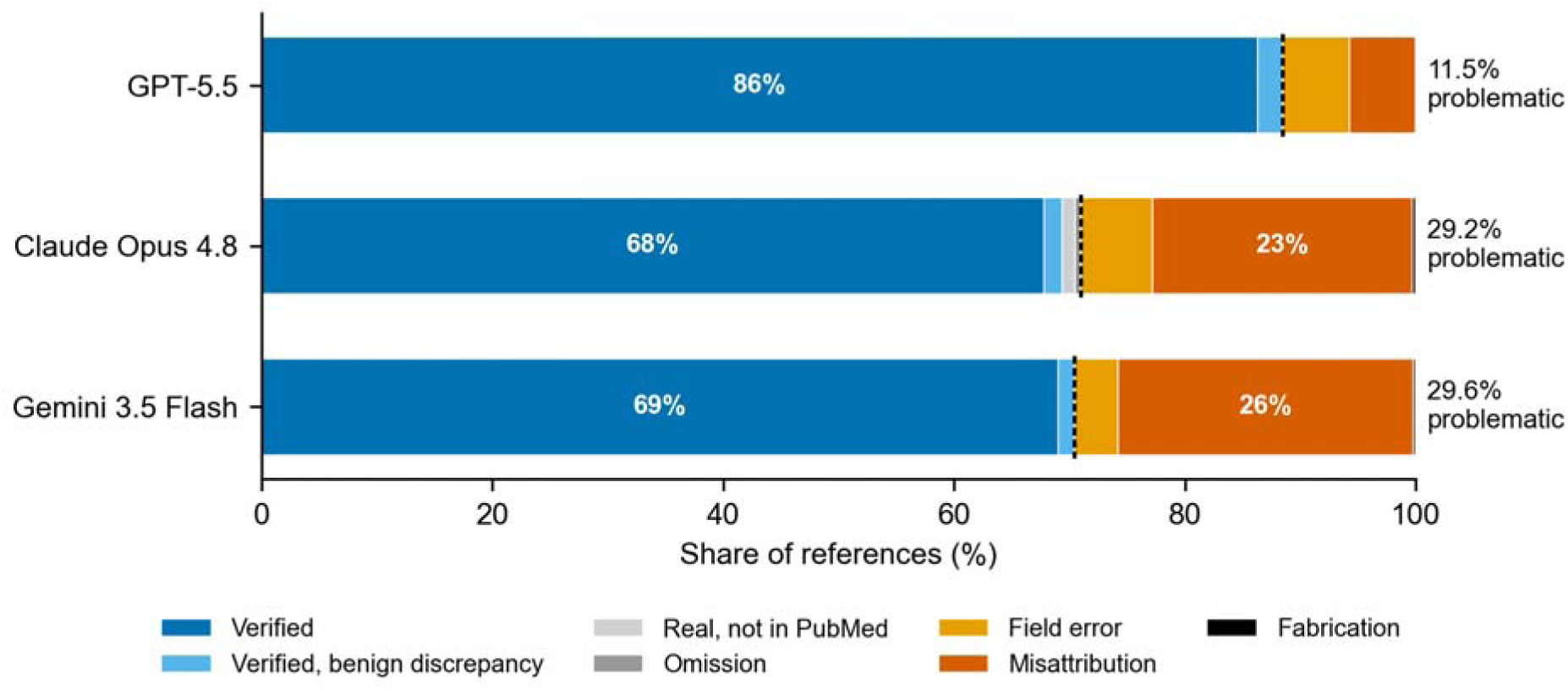
Verification outcome composition by model. Stacked bars show the share of all references generated by each model (GPT-5.5 n = 2,700, Claude Opus 4.8 n = 2,648, Gemini 3.5 Flash n = 2,702) in each of the seven verification categories. The dashed line separates non-problematic outcomes (left) from problematic outcomes (right).

**Table 2.** Verification outcomes by model.

| **Verification outcome** | **GPT-5.5**<br>**(N = 2,700)** | **Claude Opus 4.8**<br>**(N = 2,648)** | **Gemini 3.5 Flash**<br>**(N = 2,702)** |
| --- | --- | --- | --- |
| Verified, n (%) | 2,330 (86.3) | 1,796 (67.8) | 1,865 (69.0) |
| Verified benign, n (%) | 59 (2.2) | 42 (1.6) | 37 (1.4) |
| Real but not in PubMed, n (%) | 0 (0.0) | 28 (1.1) | 0 (0.0) |
| Field error, n (%) | 157 (5.8) | 164 (6.2) | 103 (3.8) |
| Misattribution, n (%) | 154 (5.7) | 597 (22.5) | 692 (25.6) |
| Fabrication, n (%) | 0 (0.0) | 7 (0.3) | 5 (0.2) |
| Omission, n (%) | 0 (0.0) | 14 (0.5) | 0 (0.0) |
| References with stated identifier, n | 2,700 | 2,634 | 2,702 |
| Problematic references, n (%)* | 311 (11.5) | 768 (29.2) | 800 (29.6) |

\*Problematic is defined as field error, misattribution, or fabrication, and its percentage uses references with a stated identifier as the denominator, the primary-outcome denominator.

### Validation of CoVe against expert adjudication

Among 270 adjudicated references, 62 (23.0%) were problematic, including 51 misattributions, 7 field errors, and 4 fabrications. CoVe returned a verdict for all 270 references and detected 60 of the 62 problematic references, corresponding to a sensitivity of 96.8% (95% confidence interval [CI] 89.0% to 99.1%) and a specificity of 98.6% (95% CI 95.8% to 99.5%) (Fig. 3). Detection was complete for misattribution and fabrication, with CoVe identifying 51 of 51 misattributions and 4 of 4 fabrications. The two missed problematic references were field errors, giving 5 of 7 field errors detected.

**Figure 3.**
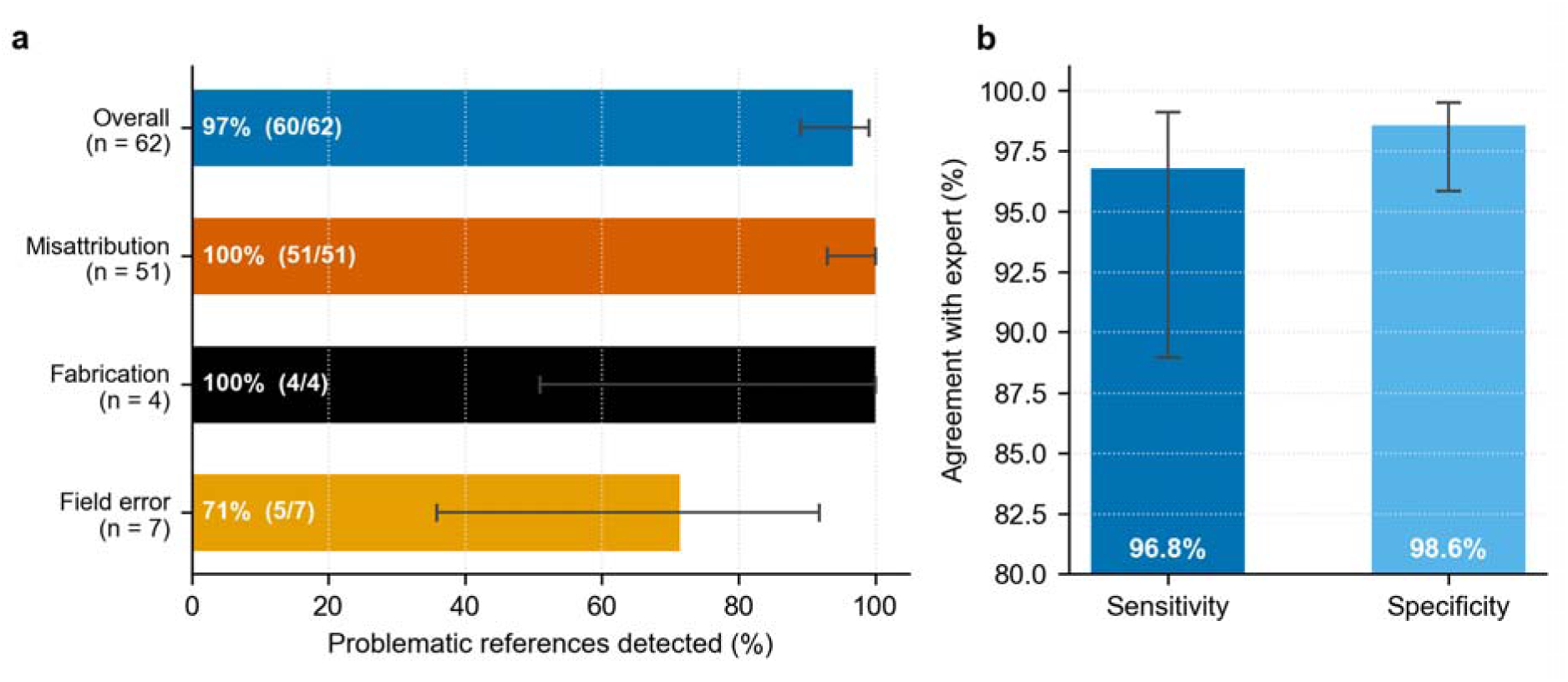
Detection of problematic references by Chain-of-Verification against expert adjudication. a, Proportion of expert-identified problematic references detected by CoVe, overall and by error type, in the 270-reference validation set (62 problematic). b, Sensitivity (60 of 62) and specificity (205 of 208) of CoVe against the expert gold standard with Wilson 95% confidence intervals. The gold standard rested on a single expert reviewer.

CoVe disagreed with the expert adjudication in five references. Two were false negatives, both field errors involving author-list discrepancies. Three were false positives in references the expert classified as real. CoVe reached this performance on a first pass, without being tuned to the validation set. Unlike a bibliographic field check, CoVe can also be extended to claim-level verification, as demonstrated in Supplementary Note 1 and Supplementary Table 10.

## Discussion

In this evaluation of three current frontier LLMs, citation reliability differed substantially by model. GPT-5.5 produced problematic references in about 12% of cases, compared with about 29% for Claude Opus 4.8 and Gemini 3.5 Flash. The dominant error was not an obviously fabricated article, but a misattributed identifier that pointed to a different real article. This finding matters for authors and editors because a misattributed identifier looks like a valid citation on inspection. Verification should therefore confirm that the identifier corresponds to the article named by the model. In the expert-adjudicated validation set, the LLM-based Chain-of-Verification approach, which verifies each citation through separate checks of its bibliographic fields, detected 60 of 62 problematic references, including all misattributions and fabrications, with specificity of 98.6%.

Earlier reports of fabricated or inaccurate citations were obtained from general-purpose chatbots, usually without integrated search tools [3–5]. Our findings refine that picture for current tool-enabled frontier models in two ways. The rate is lower for the best model, about 12% for GPT-5.5, but remains high for the others at about 29%. The nature of the error has shifted from fabrication toward misattribution, which is more difficult to detect by casual inspection. This reframes the verification problem. Because the dominant failure is a valid identifier that resolves to the wrong paper, verification must confirm that each identifier corresponds to the cited article.

The absence of a statistically significant publication-volume effect should not be interpreted as evidence that topic context is irrelevant. Linardon et al. [13] found that citation fabrication and bibliographic accuracy varied by topic familiarity and prompt specificity within mental health research. Our study tested a related but distinct construct, PubMed-indexed publication volume in cardiology, under tool-enabled generation. The numerical gradient toward fewer problematic references in higher-volume topics is consistent with the broader idea that source availability may matter, but the evidence here was not strong enough to support a definitive topic-volume effect.

The CoVe validation arm also clarifies why a Chain-of-Verification approach is well suited to citation checking. A reference is not a single fact but a linked set of claims about an article, including its title, authors, journal, year, PMID, DOI, and the statement it is used to support. Asking an LLM to judge the whole reference at once may reproduce the same error that generated the citation. CoVe instead decomposes verification into smaller questions, resolves each question against an external source, and then compares the returned values with the stated citation. In this study, that structure directly targeted the dominant failure mode, in which a real identifier was attached to the wrong article. CoVe was applied using the predefined procedure, without post hoc modification after reviewing the validation results. Its five disagreements with expert adjudication were explainable field-level discrepancies rather than failures to detect misattribution or fabrication.

The claim-level demonstration extends the same principle beyond bibliographic accuracy. A citation can be bibliographically correct and still fail to support the sentence to which it is attached. This distinction is increasingly important because recent evaluations show that medical LLM outputs may cite real sources that do not fully support their claims [17,18]. In that setting, reference verification should be understood as a stepwise process. The first step is confirming that the article exists and that the identifier belongs to the stated article. The next step is confirming that the cited article actually supports the claim made in the manuscript.

This study has several limitations. First, the study was restricted to cardiology topics and to references intended to be PubMed-indexed articles. The findings may not generalize to other specialties, nonbiomedical literature, books, guidelines, preprints, or references outside PubMed. The publication-volume gradient also did not reach statistical significance, so this finding should be interpreted cautiously. Second, the model comparison reflects the deployed API configurations available at the time of the study. Native web-search or grounding tools differed across providers, and Gemini did not expose grounding metadata, so its search behavior was inferred from the outputs rather than directly observed. Gemini was also represented by the Flash model available to the study account, which may not be directly comparable with the highest-capability models from other providers. Third, the expert reference standard was based on a single cardiologist rather than multiple independent reviewers. The CoVe validation set included 270 references, and larger independent validation samples are needed to estimate performance more precisely. The claim-level verification demonstration included only nine references and used abstracts alone, so it should be interpreted as a feasibility example rather than an estimate of claim-level error frequency. In addition, because a model from the Claude family assisted the analysis and drafting, bias toward that family cannot be excluded and is disclosed.

Current frontier LLMs vary widely in citation reliability and, even at their best, make enough errors, predominantly valid but misattributed identifiers, to require verification before clinical or scholarly use. Confirming that each identifier matches the cited article is the highest-yield safeguard, and an LLM-based Chain-of-Verification provides it at accuracy comparable to an expert.

### Use of generative artificial intelligence

The outputs under evaluation were generated by Claude Opus 4.8, GPT-5.5, and Gemini 3.5 Flash via their APIs as described above. Claude (Anthropic) was additionally used to assist code writing, the CoVe verification runs, and manuscript editing. All results were verified by the authors, who take full responsibility for the content. No LLM is listed as an author.

## Data availability

The generation prompts, the verification outcome for every one of the 8,050 references, the expert adjudication of the 270-reference validation set, and the CoVe verdicts are available in the project repository at https://github.com/JeonKH81/llm-citation-reliability and in the Supplementary Information.

## Code availability

The automated PubMed-based verification program, the CoVe run scripts, and the analysis code that reproduces all statistics, tables, and figures are available at https://github.com/JeonKH81/llm-citation-reliability.

## Acknowledgements

This study received no funding.

## Author contributions

K.-H.J. and R.S. conceived and designed the study, generated and verified the data, performed the statistical analysis, adjudicated the expert reference standard, and drafted the manuscript. J.-M.L., J.P., J.-S.K., H.-W.C. and S.-H.K. contributed to the interpretation of the data and critically revised the manuscript. All authors read and approved the final manuscript.

