## Supplementary Information for "Citation reliability of frontier large language models in medical writing and its automated verification"

### Supplementary Table 1. PubMed search strings and publication-volume counts

Publication-volume level was assigned before review generation. PubMed counts were measured once on June 18, 2026, using NCBI E-utilities esearch with one fixed query per topic.

| **Publication-volume level** | **Topic** | **PubMed query** | **Count** |
| --- | --- | --- | --- |
| Low | Transcaval access | "transcaval"[tiab] AND (access[tiab] OR TAVR[tiab] OR aortic[tiab]) | 191 |
| Low | Cardiac contractility modulation | "cardiac contractility modulation"[tiab] | 274 |
| Low | Coronary slow flow phenomenon | "coronary slow flow"[tiab] | 477 |
| Moderate | Subcutaneous implantable cardioverter defibrillator | ("subcutaneous implantable cardioverter"[tiab] OR "subcutaneous ICD"[tiab]) | 1175 |
| Moderate | Bioresorbable vascular scaffold | ("bioresorbable scaffold"[tiab] OR "bioresorbable vascular scaffold"[tiab] OR "bioresorbable stent"[tiab]) | 1389 |
| Moderate | Peripartum cardiomyopathy | "peripartum cardiomyopathy"[tiab] | 1852 |
| High | Sacubitril and valsartan | ("sacubitril"[tiab] OR "ARNI"[tiab]) | 3187 |
| High | Left atrial appendage closure | ("left atrial appendage closure"[tiab] OR "left atrial appendage occlusion"[tiab]) | 3780 |
| High | Cardiac amyloidosis | "cardiac amyloidosis"[tiab] | 4849 |

### Supplementary Table 2. Standardized generation prompt

The prompt was identical for all models, with only the topic name replaced. Each generation was run in an independent API session with the provider's native web-search or grounding function enabled. No PubMed-specific retrieval tool was supplied. Each row reproduces one block of the prompt verbatim.

| **Prompt block** | **Text** |
| --- | --- |
| Role | You are writing a narrative review for a medical journal. |
| Topic | Topic: [TOPIC] |
| Task and length | Write a concise narrative review (approximately 600-900 words) on this topic, citing exactly 30 references. Use Vancouver-style numbered citations in the text (e.g., [1], [2]). |
| Search instruction | You may use web search to find and verify the references. Cite peer-reviewed articles indexed in PubMed where possible. |
| Output format | After the review body, output the full reference list as a single JSON array inside a fenced json code block. Each element must have exactly these keys. |
| Reference keys | "index": integer, 1-30, matching the in-text citation number. "authors": full author list as cited, e.g. 'Smith J, Lee K, et al.'. "title": article title. "journal": journal name. "year": integer. "volume": volume. "issue": issue, or empty string. "pages": page range. "pmid": PubMed ID. "doi": DOI. |
| Closing instruction | Provide the PMID and DOI for each reference. Output the review body first, then the JSON block. Do not add any text after the JSON block. |

### Supplementary Table 3 | Reference-verification categories and benign rules

The automated PubMed-based check assigned each generated reference to one of seven mutually exclusive categories.

| **Category** | **Definition** | **Count in main analysis** |
| --- | --- | --- |
| Verified | Correct article and all checked bibliographic fields matched PubMed | 5991 |
| Verified benign | Correct article with only prespecified benign discrepancies | 138 |
| Real but not in PubMed | Real article confirmed through Crossref, but not indexed in PubMed | 28 |
| Field error | Correct article with at least one non-benign bibliographic field wrong | 424 |
| Misattribution | Stated PMID or DOI pointed to a different article, or the named article carried an incorrect identifier | 1443 |
| Fabrication | No corresponding article was found | 12 |
| Omission | No PMID or DOI was stated | 14 |

The primary outcome, problematic reference, was defined as field error, misattribution, or fabrication among references with a stated identifier. Verified, verified benign, and real but not in PubMed references were not counted as problematic. Omitted identifiers were reported separately and excluded from the primary denominator. Benign discrepancies were prespecified before analysis and included omission of a subtitle, a one-year difference between electronic and print publication dates, differences caused by accents or other character normalization, use of an official journal abbreviation rather than the full journal title, and an absent issue field when PubMed did not record an issue.

### Supplementary Table 4. Refusal rates in the no-tools condition

The main analysis evaluated generation with native web search or grounding enabled. A supplementary no-tools condition was used only to characterize model behavior when external search was not available. In that condition GPT-5.5 frequently refused to produce citations, especially for low-volume topics.

| **Publication-volume level** | **GPT-5.5 refusals** | **Claude Opus 4.8 refusals** | **Gemini 3.5 Flash refusals** |
| --- | --- | --- | --- |
| Low | 29 of 30 | 0 of 30 | 0 of 30 |
| Moderate | 5 of 30 | 0 of 30 | 0 of 30 |
| High | 0 of 30 | 0 of 30 | 0 of 30 |

### Supplementary Table 5. No-tools and web-search problematic-reference rates

When only non-refused no-tools outputs were considered, problematic-reference rates were higher than in the main web-search condition for all models. These results are descriptive and were not used for the primary model comparison.

| **Model** | **No-tools condition** | **Web-search condition** |
| --- | --- | --- |
| GPT-5.5 | 36.7% of 1680 references | 11.5% of 2700 references |
| Claude Opus 4.8 | 36.9% of 2671 references | 29.2% of 2634 references |
| Gemini 3.5 Flash | 66.1% of 2280 references | 29.6% of 2702 references |

For the main web-search condition, reported search-call metadata were available for GPT-5.5 and Claude Opus 4.8 but not for Gemini 3.5 Flash. GPT-5.5 reported a mean of 18.1 search calls per review and Claude Opus 4.8 a mean of 11.5 search calls per review. Gemini 3.5 Flash did not return grounding metadata, so its search use could not be directly counted.

### Supplementary Table 6. Automated-check accuracy against expert adjudication

In the 270-reference expert-adjudicated validation set, the automated PubMed-based check matched the expert reference standard after adjudication of discordant cases.

| **Model** | **Sensitivity** | **Specificity** |
| --- | --- | --- |
| GPT-5.5 | 100.0% | 100.0% |
| Claude Opus 4.8 | 100.0% | 100.0% |
| Gemini 3.5 Flash | 100.0% | 100.0% |
| Pooled | 100.0% | 100.0% |

### Supplementary Table 7. Sensitivity analysis correcting for possible automated-check misclassification

A conservative sensitivity analysis applied the pre-adjudication estimates of check performance (sensitivity 89.7%, specificity 98.1%) to the observed model-specific rates using the Rogan-Gladen correction with review-clustered bootstrap 95% confidence intervals (3,000 resamples). The model ranking was unchanged.

| **Model** | **Observed problematic-reference rate** | **Corrected rate (95% CI)** |
| --- | --- | --- |
| GPT-5.5 | 11.5% | 10.9% (9.0 to 13.0) |
| Claude Opus 4.8 | 29.2% | 31.0% (28.2 to 34.0) |
| Gemini 3.5 Flash | 29.6% | 31.5% (27.3 to 35.9) |

### Supplementary Table 8. Chain-of-Verification atomic questions and tool calls

| **Question** | **Content** | **Tool call** |
| --- | --- | --- |
| Q1 | Does the stated PMID or DOI resolve, and to which article? | PubMed record retrieval |
| Q2 | Which PMID does PubMed itself assign to the stated title and first author? | PubMed reverse search |
| Q3 | Who are the authors of the confirmed record? | PubMed record retrieval |
| Q4 | In which journal and year was the confirmed record published? | PubMed record retrieval |
| Q5 | What are the volume and pages of the confirmed record? | PubMed record retrieval |

### Supplementary Table 9. Chain-of-Verification verdict rules

| **Outcome of Q1 and Q2** | **Outcome of Q3 to Q5** | **Verdict** |
| --- | --- | --- |
| Stated identifier resolves to the article named by the model | All fields match, or only benign discrepancies | Verified |
| Stated identifier resolves to the article named by the model | At least one non-benign field wrong | Field error |
| Stated identifier resolves to a different real article, and reverse search recovers the named article | Not applicable | Misattribution |
| Stated identifier does not resolve, but reverse search recovers the named article | Not applicable | Misattribution |
| Neither the identifier nor reverse search locates the named article in PubMed or Crossref | Not applicable | Fabrication |
| Tool calls failed for the reference | Not applicable | Unverifiable |

### Supplementary Note 1. Claim-level verification feasibility demonstration

The primary validation arm evaluated verification at the level of the bibliographic record, meaning whether the cited paper exists and whether its identifiers and fields are correct. A verified record can still be cited for a statement that the paper does not actually support. This content-level error is invisible to any check that compares bibliographic fields. Chain-of-Verification extends naturally to this level through two additional questions, claim support and citation appropriateness. We therefore ran a small feasibility demonstration on nine field-verified references, one per validation review, selected in advance and blind to whether the claim would hold, as described in the Methods.

Among the nine verified references, five claims were fully supported by the cited abstract, two could not be confirmed from the abstract alone, one showed a minor numerical discrepancy, and one directly contradicted the cited paper (Supplementary Table 10).

The clearest content error involved a real, correctly identified paper. Claude Opus 4.8 cited a subgroup analysis of a cardiac contractility modulation trial (PMID 21872139) for the statement that patients with a left ventricular ejection fraction between 35% and 45% benefited the most and that those below 25% benefited less. According to the cited paper, the parent trial enrolled only patients with an ejection fraction of 35% or less, and the responsive subgroup was defined by an ejection fraction of 25% or more, meaning 25% to 35%. No 35% to 45% subgroup exists in the trial. The bibliographic record was correct, so field-level verification classified the reference as correct. Only the claim-level check detected that the cited paper did not support the statement. A second, milder discrepancy involved a bioresorbable scaffold meta-analysis (PMID 26597771). The direction of the claim was correct, but the patient counts and event rates cited in the review did not match those in the cited abstract, suggesting conflation with a different pooled analysis.

This demonstration shows that claim-level verification catches a class of error that bibliographic verification cannot, namely a real paper cited for a statement it does not support. It covers nine references and is not powered for rate estimation, judgments rested on the abstract alone, and a single reviewer performed the assessment. Claim-level verification should be evaluated formally in future work with full-text access, multiple reviewers, and a larger sample.

### Supplementary Table 10. Claim-level verification of nine field-verified references

| **Review** | **Model** | **Cited article (PMID)** | **Claim support** | **Severity** |
| --- | --- | --- | --- | --- |
| R1 | Gemini 3.5 Flash | 2022 AHA/ACC/HFSA heart failure guideline (35363499) | Not in abstract | None |
| R2 | Gemini 3.5 Flash | PARADIGM-HF (25176015) | Supported | None |
| R3 | GPT-5.5 | BIOSOLVE-I DREAMS (23332165) | Supported | None |
| R4 | Gemini 3.5 Flash | EHRA/EAPCI LAAO consensus (31504441) | Unverifiable, no abstract indexed | None |
| R5 | Claude Opus 4.8 | CCM subgroup analysis (21872139) | Contradicted | High |
| R6 | GPT-5.5 | NHLBI transcaval TAVR (27989885) | Supported | None |
| R7 | Claude Opus 4.8 | ATLAS S-ICD trial (36343346) | Supported | None |
| R8 | GPT-5.5 | Transcaval versus transaxillary TAVR (35512920) | Supported | None |
| R9 | Claude Opus 4.8 | BVS meta-analysis (26597771) | Partially supported | Low |
